# Human papillomavirus vaccination at the national level in Tunisia: a cost-effectiveness analysis using a comparative modeling study

**DOI:** 10.1101/2024.09.17.24313756

**Authors:** Oumaima Laraj, Beya Benzina, Ahlem Gzara, Amira Kebir, Kaja Abbas, Slimane Ben Miled

## Abstract

Cervical cancer is one of the most prevalent cancer diseases in women caused by persistent infection with one of 13 sexually transmitted high-risk human papillomavirus (HPV) types. Vaccination can significantly reduce the prevalence of this burden in low-middle income countries. However, HPV vaccination is not included in the Tunisian immunization program. Since the economic evaluation of HPV vaccines is crucial to inform public-health decisions, the World Health Organization (WHO) recommends that a cost-effectiveness analysis of HPV vaccination is conducted before nationwide introduction. This study aimed to conduct a cost-effectiveness analysis of incorporating different HPV vaccines into the national immunisation schedule in Tunisia. The potential health and economic impacts of human papillomavirus (HPV) vaccination were evaluated through comparative modelling analysis using two published static models (UNIVAC and Papillomavirus Rapid Interface for Modelling and Economics (PRIME)). Academic literature and anecdotal evidence were included on the demographic variables, cervical cancer incidence and mortality, treatment costs, vaccine delivery costs and other model parameters.

The cost of vaccination, treatment costs saved, net costs, cases and deaths averted, life years saved, disability-adjusted life years (DALYs) prevented, and incremental cost-effectiveness ratios were predicted and reported as primary outcomes. The incremental cost-effectiveness ratios (ICERs) were estimated per disability-adjusted life years (DALYs) averted using the cost-effectiveness threshold (CET) defined by the World Health Organisation (WHO). All HPV vaccines were very cost effective (with every disability-adjusted life-year averted costing less than the cost-effectiveness threshold). The analyses were done from a health system and societal perspective. Despite model differences, the PRIME and UNIVAC models yielded similar vaccine-impact estimates.

## 1 Introduction

In November 2020, the World Health Organization (WHO) launched a global initiative to accelerate the elimination of cervical cancer as a public health issue. This initiative focuses on three key strategies: vaccination, screening, and treatment [37]. Cervical cancer, like many cancers in women, is primarily caused by human papillomavirus (HPV) infection, which accounts for over 90% of cases [8]. HPV is a DNA virus with several genotypes, some of which are categorized as high-risk due to their cancer-causing potential. Specifically, HPV genotypes 16 and 18 are responsible for about 70% of global cervical cancer cases, while other high-risk genotypes include HPV 31, 33, 45, 52, and 58 [9].

Effective prevention strategies include vaccination against the most common HPV genotypes and regular screening through cervical cytology and HPV testing. The WHO’s goal is to achieve 90% HPV vaccination coverage for girls by age 15, screen 70% of women, and ensure that 90% of those diagnosed receive treatment (90-70-90). Thus, HPV vaccination is a crucial component of cervical cancer prevention efforts.

In Tunisia, HPV genotypes 16 and 18 are the most prevalent and significantly contribute to cervical cancer cases. Other high-risk genotypes such as HPV 31, 45, 51, and 56 are also common.

In January 2024, Tunisia’s Ministry of Health announced plans to incorporate the HPV vaccine into the national school vaccination program starting in 2025 for girls in the 6th year of primary school [30]. With an approximately 97% vaccination coverage in schools and a 92% enrollment rate, Tunisia is well-positioned for widespread HPV vaccination.

Mathematical models are essential for studying the spread and control of HPV, as they simulate its progression in populations using demographic, epidemiological, and clinical data [28, 29]. These models estimate the number of cervical cancer cases, deaths averted, and incremental costs per Disability-Adjusted Life Year (DALY), a key public health measure that combines years lost to premature death and years lived with HPV-related disability. For our analysis in Tunisia, we used and compared the outputs of the PRIME model (Papillomavirus Rapid Interface for Modeling and Economics) [3] and the UNIVAC model [11].

Given budget constraints, it is important for decision-makers to conduct an epidemiological and economic analysis to determine the most suitable prevention modalities for Tunisia, including the choice of vaccine. The national technical vaccination committee initially decided to follow the WHO’s strategy of using a single dose of the quadrivalent vaccine [10]. The negotiated price of this vaccine is $17 USD, including all related charges. However, bivalent vaccines (Cervarix and Cecolin) might offer a better cost-effectiveness ratio at a lower prices of $12 USD and $5 USD, respectively, including charges, prior to any negotiation.

The primary objective of this study is to evaluate the health and economic impacts of HPV vaccination for 12-year-old girls in Tunisia, considering various vaccine options, their effectiveness, costs, and cost-effectiveness. Specifically, the study aims to assess the health benefits of different HPV vaccines (CECOLIN, CERVARIX, GARDASIL-4, and GARDASIL-9) by estimating the reduction in cervical cancer cases, deaths, and disability-adjusted life years (DALYs) over the lifetime of a birth cohort vaccinated in 2025, both with and without cross-protection. Additionally, we will analyze the economic outcomes of each vaccine option, including discounted implementation and treatment costs, and evaluate the cost-effectiveness from both governmental and societal perspectives. These analyses, obtained from the UNIVAC and PRIME models, will help determine the consistency of health and economic outcomes and identify variations due to differences in modeling approaches.

The paper is organized as follows: we begin by summarizing the materials and methods and providing the data used in disease burden, healthcare costs, and vaccination parameters in the appendix. Section 3 presents a comparative analysis of vaccine impact projections using the PRIME and UNIVAC models, evaluates the projected cost-effectiveness and budget impact of incorporating the HPV vaccine into Tunisia’s national immunization program, and conducts sensitivity analyses to assess the impact of parameter uncertainty on model results. Finally, we identify the most effective approach for reducing the burden of cervical cancer in Tunisia, discuss the strengths and limitations of both models, and present our conclusions and future perspectives.

## 2 Methods

### 2.1 Modeling approach

We adapted the PRIME and UNIVAC models to project the health and economic impact of HPV vaccination at a national level in Tunisia. Both the PRIME and UNIVAC models are static multi-cohort, proportional impact models used to estimate the impact of HPV vaccination on cervical cancer cases and deaths. The UNIVAC model uses United Nations (2019 revision) population estimates and evaluates catch-up campaigns, stratified cervical cancer cases by stage, and hospitalizations, while the WHO-supported PRIME model focuses on the cost-effectiveness of vaccinating females before sexual debut, utilizing country-specific data and customizable inputs. Both models assume no cross-protection or indirect effects and maintain constant age-specific cervical cancer incidence among unvaccinated women, making their estimates conservative. Table 6 in appendix outlines the similarities and differences between these models.

We evaluate the vaccination of 12-year-old girls in 2025 analyzing a cohort of girls born in 2013. This group of girls was followed up to the age of 100 years. Input data on birth and vaccination cohort sizes were obtained from the Tunisian National Institute of Statistics [27]. We estimated the number of cases, deaths, and disability-adjusted life years (DALYs) with and without vaccination. Burden estimates were aggregated over the lifetimes of each cohort of vaccinated girls. The direct impact of vaccination is calculated for each year of age by multiplying vaccine coverage by vaccine efficacy, adjusted for the HPV type distribution and the assumed efficacy of each vaccine product against each HPV type. The model also estimates the costs of the HPV vaccination program and healthcare costs, with and without vaccination.

Model inputs related to vaccine aspects (*e.g* efficacy, program costs, and delivery expenses), as well as cervical cancer considerations, including disease burden and treatment costs, were sourced from a combination of published local and global references [13, 32, 16] and insights provided by local stakeholders [25].

To eliminate the effect of the time value of money, all future costs and health benefits were discounted at the rate of 3 % over a lifetime time horizon, based on the recommendation from WHO guidelines on health economics [2] for immunization programs. All costs represent 2024 USD $. We calculated the probability of vaccine being cost-effective over a range of alternative possible WTP thresholds up to 0.3 times the national GDP per capita (US $ 3747 in the year 2024) [27] as Tunisia does not have a strict willingness-to-pay (WTP) threshold for determining the cost-effectiveness of an intervention.

We are approaching the problem from a societal and governmental perspective. From the perspective of the health and societal system, we compared the final costs and health effects of the two strategies of not vaccinating and vaccinating of the target age group. The results of the economic evaluation were expressed by incremental cost-effectiveness ratios (ICER), and ICER indicators that were constructed based on the disability-adjusted life years (DALYs), were reported for the two modeling approaches. The evaluation assumed that the target population had not been infected with HPV prior to vaccination. The primary outcome measure is the cost (US$) per DALY averted, accounting for all costs and benefits aggregated over the cohort of vaccinated girls (2025).

### 2.2 Disease burden

We used age-specific rates of cervical cancer cases and deaths estimated for Tunisia from the global database of GLOBOCAN 2022 [13] and assumed these rates related to local, regional and distant stages [12] would remain constant over time in the absence of vaccination (Figure 1). For the proportion of cervical cancer that is attributed to the HPV genotype targeted by the vaccines (*e.g.,* HPV 16/18 and HPV 16/18, 31, 33, 45, 52, and 58), we used estimates provided by [16]. Inputs for disease burden are summarized in Table 7.

**Figure 1:**
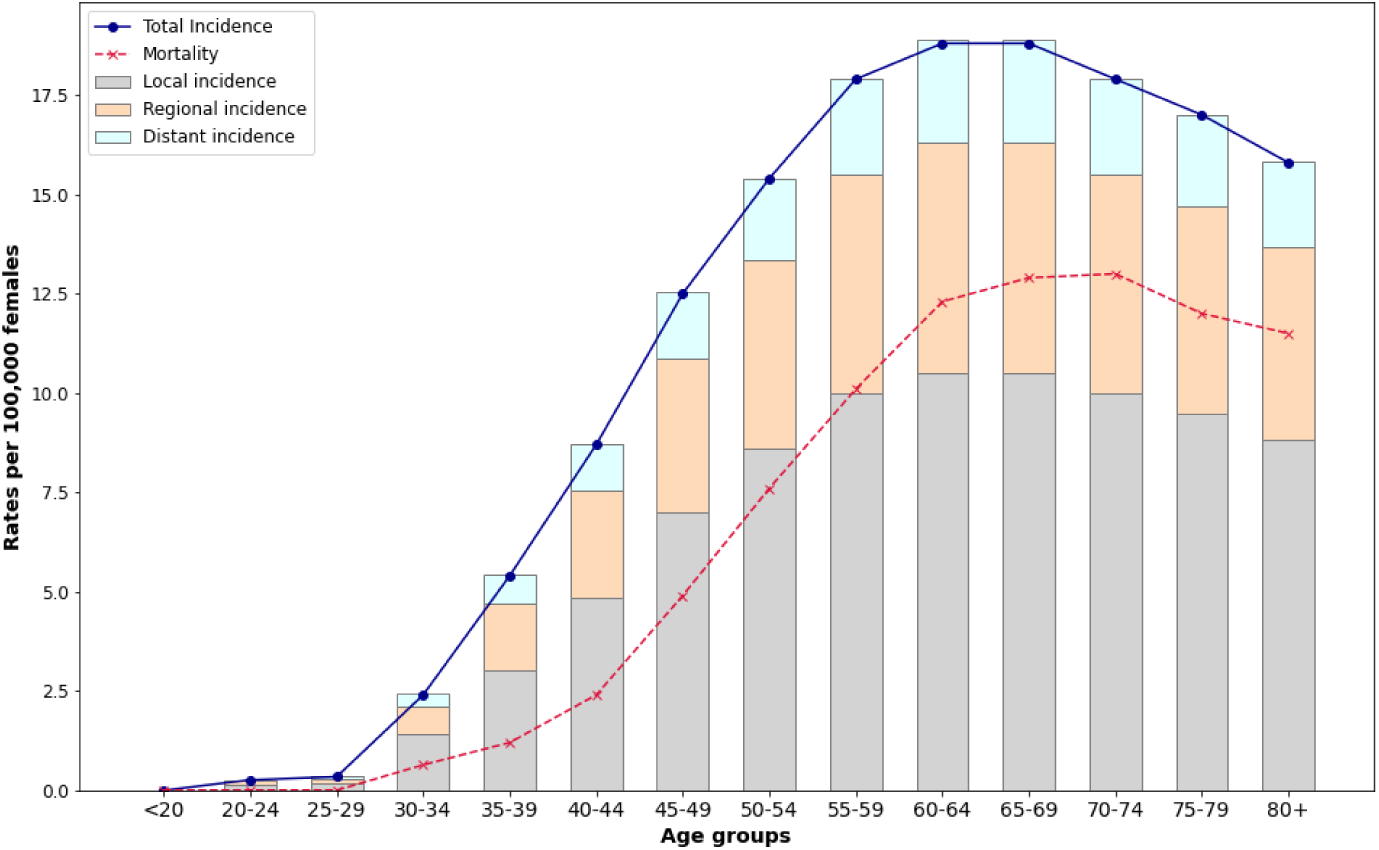
Age-specific cervical cancer burden by stage in Tunisia (2022).

The disability weights and durations for the different phases of cervical cancer are used in estimating the years of life lost due to disability. Disability weights are evaluated for the different phases of cervical cancer: diagnosis and primary treatment phase, non-terminal sequelae phase and terminal phase, based on the Global Burden of Disease (GBD) studies [22]. In the UNIVAC model, we assumed that cases were distributed into local, regional, and distant cancer categories, using the International Federation of Gynecology and Obstetrics (FIGO) staging system and information from published local studies [12]. Age-specific disease incidence, mortality and cancer distribution are reported in Figure 1. In addition, disability weights to represent time lost while living with local, regional, and distant cancer were taken from [31]. Average five-year survival rates were based on a recent report of cervical cancer survival in Tunisia from National Cancer Institute in 2023 [25].

### 2.3 Healthcare costs

We assumed that all women included in the GLOBOCAN incidence rates [13] would be diagnosed and receive treatment, so the average cost of cervical cancer treatment was applied to them.

Direct treatment-related costs were derived from an existing cost study in the Salah Azaiez Institute [25] that estimated the stage-specific treatment costs for cervical cancer in Tunisia. Each stage of cancer classification required different medical interventions including clinical, biological, radiological, and pharmaceutic resources. The economic direct cost of treating the various stages of cervical cancer in Tunisia in 2023 ranged between US $ 532 and US $ 2603, depending on the stage of the disease [25]. This includes costs related to diagnosis, staging, surgery (simple/radical hysterectomy), chemotherapy, radiotherapy, and palliative care. In PRIME, the cancer treatment cost per episode over lifetime was calculated as the average cost of treatment for the different procedures related to three stages. Inputs for healthcare costs are summarized in Table 8 (see appendix). In contrast, direct medical costs of cervical cancer treatment by stage is considered in the UNIVAC model. The FIGO (International Federation of Gynecology and Obstetrics) staging system, ranging from early localized stages (IA and IB) to more advanced stages (II, III, IVA) and distant metastases (IVB), is used to classify the extent of cervical cancer. Costs associated with these stages vary significantly, reflecting the complexity and intensity of required interventions. For FIGO Stage IA, involving very early and localized cancer, the cost is approximately $550, covering the initial gynecological examination, inpatient stay, and necessary pre-operative tests. In contrast, Stages IB1 and IB2, which involve more extensive but still localized cancer, have a mean cost of $585, including laparoscopy, curettage, radical hysterectomy or trachelectomy, and possible hospital stays. More advanced stages, such as IB3, II, IIIA, IIIB, and IVA, which require complex treatments like PET scans, radiotherapy, chemotherapy, and brachytherapy, incur significantly higher costs, averaging $2603. Stage IIIC, characterized by lymph node involvement, also requires intensive treatment with a mean cost of $1800. For Stage IVB, involving distant metastasis, the cost averages $750, covering palliative chemotherapy and related hospital care. Thus, we estimated the overall costs of cervical cancer treatment at $2603 for local stages, $1800 for regional stages, and $750 for distant stages, with an assumed average cancer treatment cost of $2445.14 in the PRIME model.

Treatment costs from a societal perspective was evaluated for the cost-effectiveness analysis. These costs included both direct medical costs and indirect costs (opportunity costs of women’s time for procedures). Indirect treatment costs, limited to productivity loss due to ill health, include convalescence if they had hospital stays or recovery time, time waiting for test results and time costs associated with travel to/from hospital visits. Productivity loss related to absenteeism was derived using the human capital approach; the total number of workdays lost multiplied by the average daily wage of a cervical cancer patient. Due to data limitations, The number of days lost due to productivity was derived from a previous micro-costing study conducted in Vietnam [35]. For productivity loss due to illness, we applied the Tunisia 2023 minimum wage, reported as 30 Tunisian dinars corresponding to 9.68 $ per day. Thus, avearge cancer treatment cost from a societal perspective is the sum of average cancer treatment cost from a healthcare perspective and productivity loss (estimated to be USD$ 1452 for 150 days of absence due to ill health).

### 2.4 Vaccination scenarios and related parameters

Plausible strategies for introducing the HPV vaccine in Tunisia were developed with input from key stakeholders, including the Tunisian Ministry of Health. The modeling assumed that vaccination would start in 2025, using a school-based delivery strategy with an expected coverage of 87%. We considered four highly effective and safe HPV vaccines currently available worldwide: **Cervarix bivalent** (Glaxo-SmithKline Biologicals, Belgium), **Cecolin bivalent** (Xiamen Innovax Biotech Co, China), **Gardasil quadrivalent** (Merck & Co., USA), and **Gardasil-9 nonavalent** (Merck & Co., USA). The bivalent vaccines target HPV types 16 and 18, the quadrivalent targets types 16, 18, 6, and 11, and the nonavalent vaccine covers additional high-risk strains (e.g., HPV 31, 33, 45, 52, 58) not included in the bivalent or quadrivalent vaccines.

Our analysis compared these vaccines against no vaccination (with no changes to existing cervical cancer screening and treatment strategies) and against each other. For our central estimates, we assumed a single-dose administration for each vaccine to a cohort of 12-year-old girls. Tunisia’s recommended school-based model is based on an 87% coverage rate. We evaluated the impact of HPV vaccination for a single cohort of 12-year-old girls in 2025, assigning protection rates of 90-98% against genotypes directly targeted by the vaccines. Specifically, we assumed 97% protection for HPV-16 and 18 with CECOLIN [24], 98% efficacy against HPV-16/18/6/11 with GARDASIL-4 [6], and 96.7% protection against HPV-16/18/31/33/45/52/58 with GARDASIL-9 [7], all for a single-dose vaccination schedule. Efficacy estimates for CERVARIX were taken from [18]. We evaluated three strategies: single-dose nonavalent vaccines at the best negotiated price, single-dose quadrivalent vaccines at the manufacturer’s listed price, and single-dose bivalent vaccines at different listed prices for CECOLIN and CERVARIX. The **baseline scenario** was no vaccination, representing the current state.

#### Vaccine impact calculations

The HPV type distribution in Tunisia was taken from estimates identified among invasive cervical cancer cases, reported by the HPV Information Center [16]. The top three prevalent HPV types were 16 (61 %), 18 (8.5 %) and 45 (5 %).

Studies have shown that one dose of HPV vaccination could provide similar benefits to two doses [14] and that vaccines offer some level of cross-protection against genotypes not covered by the vaccine [24, 6]. In our analyses, we also accounted for this cross-protection against non-vaccine types. We relied on evidence from a systematic review to estimate the degree of cross-protection offered by HPV vaccines against various genotypes [5]. In cases where multiple commercial brands of the same type of vaccine (bivalent, quadrivalent, or nonavalent) were mentioned, we averaged the degree of protection across these brands.

There is uncertainty about the scale of cross-protection to non-vaccine types that might be associated with each of the four vaccine products, so weighted efficacy values were derived by multiplying the efficacy assumed for each HPV type by the proportion of cervical cancers caused by each type in Tunisia. Based on these interpretations, we adjusted the cross-protection matrix of vaccines to reflect these observations. This updated matrix was used to estimate the effectiveness of vaccines against the HPV genotypes. The overall weighted efficacies of the four products (CECOLIN, CERVARIX, GARDASIL, GARDASIL-9) were estimated to be 69 % [24], 68 % [18], 67 % [6], and 85 %, respectively, without cross-protection. For CERVARIX, we assumed there could be cross-protective efficacy against types 31, 33, 45, 51, 52, and 56 based on a study by Wheeler et al. [39]. The influential cross-protection assumptions for GARDASIL-4 were taken from a study by Brown et al. [5] and was used with cross-protective efficacy against type 31. Additionally, the analysis of cross-protection was performed only for bivalent and quadrivalent vaccines, as we didn’t find documentation on the evaluation of the nonavalent vaccine, introduced in 2014. Figure 2 provides a detailed presentation of the cross-protection matrix of vaccines. Due to the similarity in efficacy with and without cross-protection for CECOLIN, GARDASIL-4 and GARDASIL-9, we restricted our primary analysis to five scenarios. The first four scenarios assumed no cross protection for each product. We then ran two additional scenarios for CECOLIN and CERVARIX with cross-protection.

**Figure 2:**
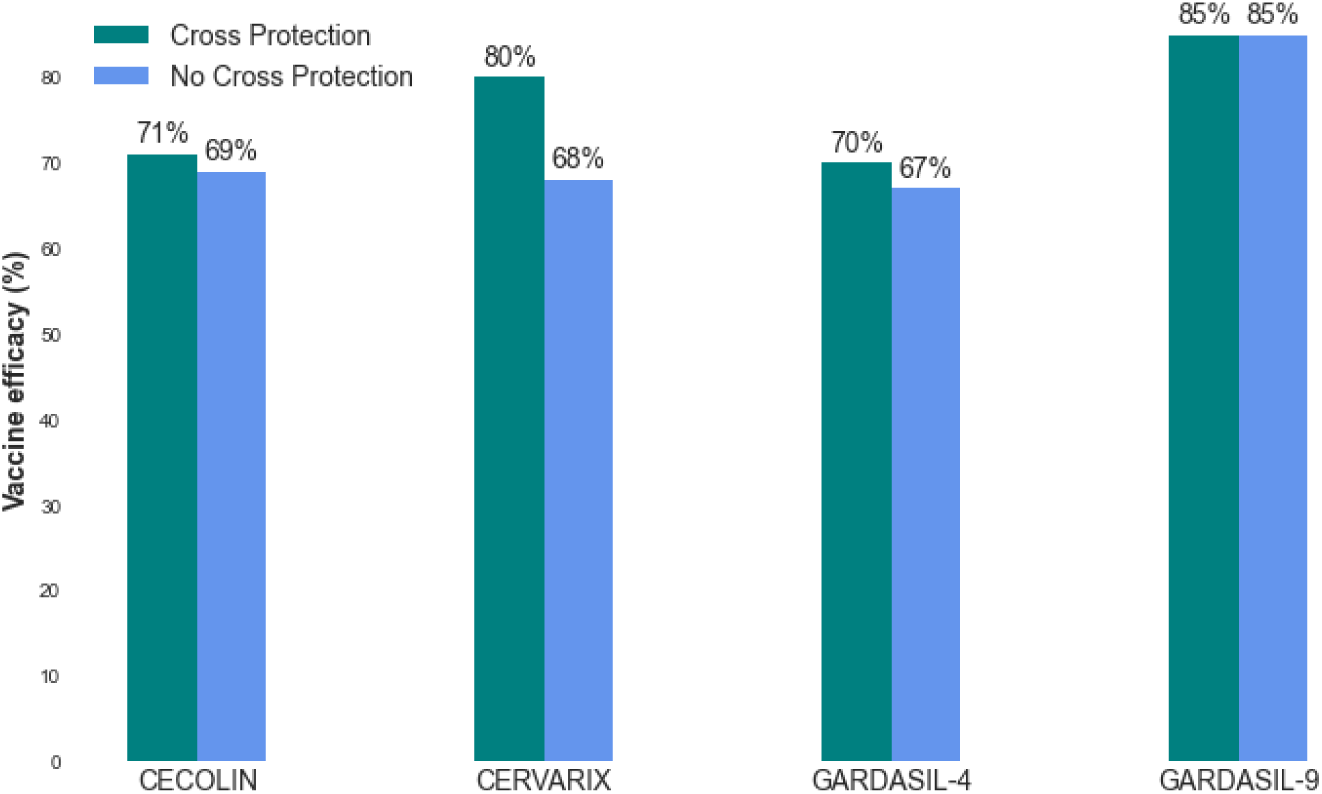
Weighted vaccine efficacy of one dose against cervical cancer cases and deaths in Tunisia with and without cross-protection.

#### Vaccine program costs

Tunisia is not eligible for vaccine financial support from Gavi. Our cost perspective included the full cost of the vaccine program borne by the government. Input data for vaccine program costs are summarized in Table 9 in the appendix, and include the costs of the vaccines, syringes, and safety boxes together with the costs of international delivery and other supplies associated with the delivery strategy (e.g, additional staff time, training, cold-chain capacity etc). As Tunisia is not eligible for receiving support through the GAVI mechanism, the prices of self-procurement of CECOLIN, CERVARIX, and GARDASIL-4 were evaluated at 3.65 $, 10.25 $ and 14.14 $, respectively [32]. GARDASIL-9 is not yet supported from GAVI, thus, we assumed a price of $25.00 per dose based on the lowest negotiated price for a non-Gavi country according to the MI4A/V3P vaccine purchase data [38]. Self-financing countries initially receive either a lump sum from Gavi vaccine introduction grants (VIGs) to subsidize the costs of HPV vaccination delivery, or support for the vaccine prices to facilitate the initial introduction of vaccination. Tunisia will receive support from Gavi, which will cover 50% of the cost of the vaccine to be procured in the first year. Additionally, funding for medical staff training and other incremental costs associated with the delivery strategy will be provided. International handling fees and costs for other supplies (syringes and safety boxes) were based on data reported from UNICEF Supply Division [36], while the delivery fee was sourced from Vodicka et al. [17]. These fees are obtained as a percentage of the dose price. We assumed a 3% international handling fee including transport and logistics, a 5% vaccine wastage for vaccines available in a one-dose vial presentation (CECOLIN, GARDASIL-4 and GARDASIL-9) and a two-dose vial for CERVARIX. We assumed further a 10% international delivery fee to cover the cost of insurance, customs duties and taxes. Prices for syringes were evaluated at 0.07 $ per dose and 1.30 $ per box (with 100 syringes per safety box), respectively. The incremental health system cost per dose was estimated based on the HPV introduction plan budget made by the Tunisian government. In PRIME model, a total vaccine delivery cost per dose was calculated based on all these incremental costs.

### 2.5 Sensitive and uncertainty analysis

Sensitivity analysis was performed to evaluate the robustness of the model results. We conducted for each vaccine, a probabilistic sensitivity analysis to assess the impact of combined parameter uncertainty on the cost-effectiveness ratios. We ran separate PSAs for each vaccine product without cross-protection and one additional scenario for CERVARIX with cross-protection (1000 runs per scenario). All parameters were varied simultaneously with random draws from their plausible ranges. Prices were assumed to be fixed within the PSA and 95% uncertainty intervals was assumed to represent the 2.5*th* and 97.5*th* percentiles of probabilistic simulations. For each probabilistic simulation, parameters were drawn from a distribution with a mean equal to the point estimate and range equal to the low and high values of the uncertainty range. In the absence of information about the shape of each distribution, the low, mid and high values for each input parameter were assumed to represent the mode and range within a series of PERT-Beta distributions. PSA results were represented as clouds on a cost-effectiveness plane and used to estimate the probability that each vaccine would be cost-effective at different WTP thresholds (cost-effectiveness acceptability curves).

## 3 Results

### 3.1 Health benefits

Vaccinating 12-year-old girls in 2025 involves vaccinating a single birth cohort of girls (born in 2013). Without HPV vaccination in Tunisia, UNIVAC estimates there could be around 788 cases, 462 deaths attributed to cervical cancer and 2495 DALYs (discounted) over the lifetime of this birth cohort.

Without cross-protection, CECOLIN, CERVARIX, and GARDASIL-4 would each have a similar projected health impact (around 60 % reduction in cervical cancer cases and deaths) during the lifetime of the vaccinated cohort. The impact of GARDASIL-9 is estimated to be around 74 % (Table 1).

**Table 1:** Lifetime health effects of each vaccine option (Bivalents: CECOLIN, and CERVARIX, quadrivalent: GARDASIL-4 and nonavalent: GARDASIL-9) compared to no vaccine and to each other in the UNIVAC model for Tunisian girls aged 12 years vaccinated in 2025 without cross protection.

| No cross protection | No vaccine | CECOLIN | CERVARIX | GARDASIL-4 | GARDASIL-9 |
| --- | --- | --- | --- | --- | --- |
| **Health outcomes** |  |  |  |  |  |
| Cervical cancer cases (local) | 440 | 176 | 180 | 183 | 115 |
| Cervical cancer cases (regional) | 243 | 97 | 99 | 101 | 63 |
| Cervical cancer cases (distant) | 106 | 42 | 43 | 44 | 28 |
| Cervical cancer cases with treatment | 788 | 315 | 322 | 329 | 205 |
| Cervical cancer deaths | 462 | 185 | 189 | 193 | 120 |
| DALYs (discounted) | 2495 | 997 | 1019 | 1041 | 650 |
| **Differences (comparator = no vaccine)** |  |  |  |  |  |
| Cervical cancer cases (local) | - | 264 | 260 | 256 | 325 |
| Cervical cancer cases (regional) | - | 146 | 144 | 142 | 180 |
| Cervical cancer cases (distant) | - | 63 | 62 | 62 | 78 |
| Cervical cancer cases with treatment | - | 473 | 466 | 459 | 583 |
| Cervical cancer deaths | - | 277 | 273 | 269 | 342 |
| DALYs (discounted) | - | 1498 | 1476 | 1454 | 1845 |
| Reduction in disease burden (%) | 0 % | 60 % | 59.2 % | 58.3 % | 74 % |

In scenarios with cross-protection, CECOLIN and GARDASIL-4 would be expected to avert 62% and 61 % of cervical cancer cases and deaths, respectively. In contrast, the health impact of CERVARIX increased to around 70 % and had substantially more health benefits than the other two products. Equivalent estimates for GARDASIL-9 were 74% (Table 2).

**Table 2:** Lifetime health effects of each vaccine option (Bivalents: CECOLIN and CERVARIX, quadrivalent: GARDASIL-4 and nonavalent: GARDASIL-9) compared to no vaccine and to each other in the UNIVAC model for Tunisian girls aged 12 years vaccinated in 2025 with cross protection.

| Cross protection | No vaccine | CECOLIN | CERVARIX | GARDASIL-4 | GARDASIL-9 |
| --- | --- | --- | --- | --- | --- |
| **Health outcomes** |  |  |  |  |  |
| Cervical cancer cases (local) | 440 | 168 | 134 | 172 | 115 |
| Cervical cancer cases (regional) | 243 | 93 | 74 | 95 | 63 |
| Cervical cancer cases (distant) | 106 | 40 | 32 | 41 | 28 |
| Cervical cancer cases with treatment | 788 | 301 | 240 | 308 | 205 |
| Cervical cancer deaths | 462 | 177 | 140 | 181 | 120 |
| DALYs (discounted) | 2495 | 954 | 758 | 976 | 650 |
| **Differences (comparator = no vaccine)** |  |  |  |  |  |
| Cervical cancer cases (local) | - | 272 | 306 | 268 | 325 |
| Cervical cancer cases (regional) | - | 150 | 169 | 148 | 180 |
| Cervical cancer cases (distant) | - | 65 | 74 | 64 | 78 |
| Cervical cancer cases with treatment | - | 487 | 549 | 480 | 583 |
| Cervical cancer deaths | - | 285 | 322 | 281 | 342 |
| DALYs (discounted) | - | 1541 | 1737 | 1519 | 1845 |
| Reduction in disease burden (%) | 0 % | 61.8 % | 69.7 % | 60.9 % | 74 % |

Over the same period, the PRIME model estimates there could be approximately 608 cases, 338 deaths and 1621 DALYs (discounted) attributed to cervical cancer. The UNIVAC model projects a higher number of cervical cancer cases and deaths averted than the PRIME model by 23% and 27%, respectively (Figure 3) assuming no cross protection. Specifically, the range of the potential health impact of HPV vaccination, in terms of the number of cervical cancer cases averted among girls vaccinated in 2025, is as follows: 364 to 473 for CECOLIN, 359 to 466 for CERVARIX, 354 to 459 for GARDASIL-4, and 449 to 583 for GARDASIL-9. Similarly, as both the years lived with disability for the estimated cervical cancer cases and the years of life lost for the estimated cervical cancer deaths contribute to the estimates of DALYs averted by the HPV vaccine, the UNIVAC model estimated 26% more DALYs averted than the PRIME model. However, the differences between the model in terms of the projected health outcomes of vaccination impact assuming no cross protection are slighty different by less than 1% (Figure 4a).

**Figure 3:**
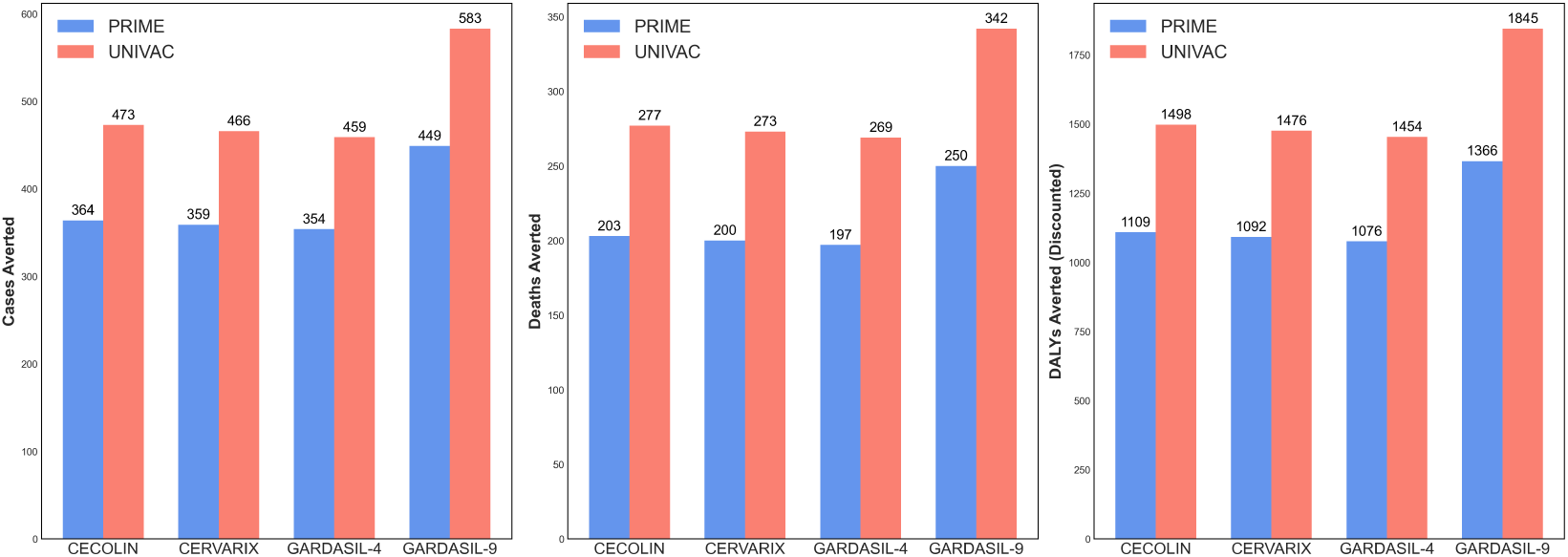
Cervical cancer cases, deaths and DALYs averted by the four products (CECOLIN, CERVARIX, GARDASIL-4 AND GARDASIL-9) among girls vaccinated in 2025 over lifetime using PRIME and UNIVAC models without cross protection.

**Figure 4:**
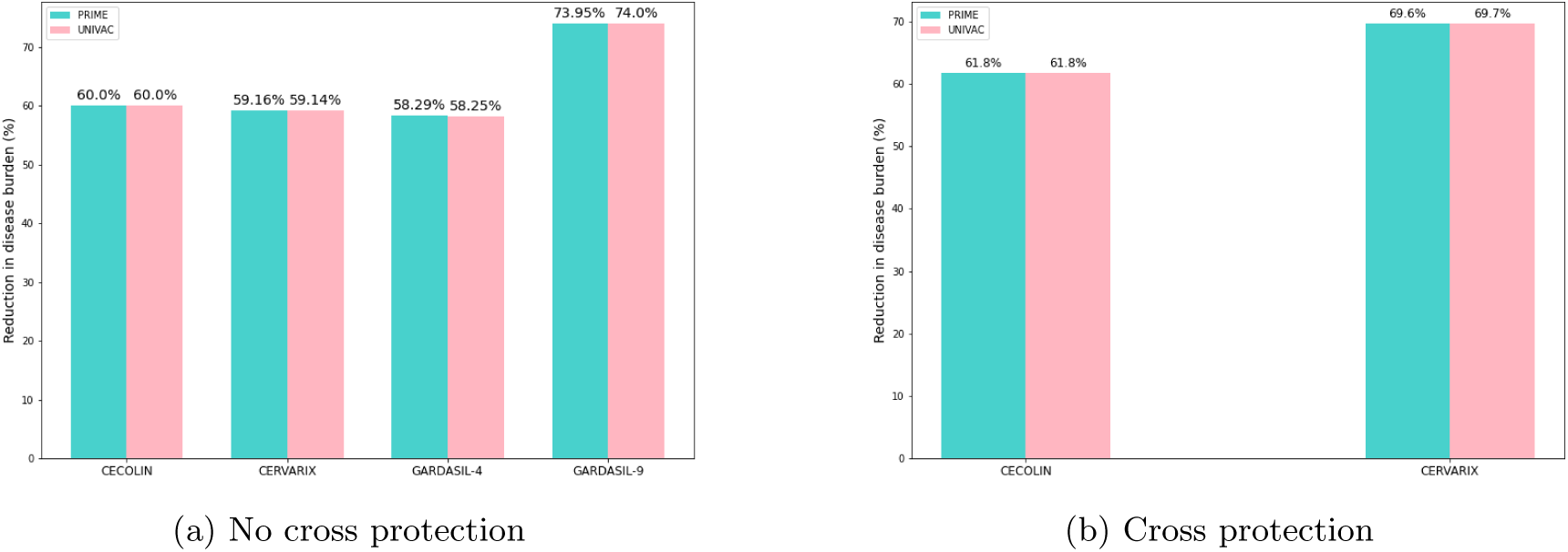
Vaccination impact in terms of reduction in disease burden among girls vaccinated in 2025 over lifetime for the four products (CECOLIN, CERVARIX, GARDASIL-4 and GARDASIL-9) using PRIME and UNIVAC models.

We assumed that CECOLIN would have approximately the same cross-protection as GARDASIL-4, and no cross-protection was assumed for GARDASIL-9. In scenarios with cross-protection, CECOLIN could prevent around 62% of cervical cancer cases and deaths, while CERVARIX could avert around 70% using both PRIME and UNIVAC models (Figure 4b).

The main difference in the projected health outcomes of vaccination impact (estimates for cases, deaths, and DALYs averted) between the two models may be due to variations in population demography and age-specific life expectancy. As cervical cancer deaths are directly estimated from cervical cancer cases, the mortality estimation approach: stage-specific (UNIVAC) versus age-specific (PRIME) is likewise the main driver of the differences in the vaccination impact. However, the reduction in disease burden by vaccination compared to the baseline scenario of no vaccination, using the two different models with and without cross protection, is not strenght and differs only slightly (Figure 4)

### 3.2 Economic outcomes

A single year of routine one-dose HPV vaccination requires substantial upfront investments in vaccine procurement and delivery. However, these costs are offset in the long term by the reduction in future cancer cases. The discounted costs of implementing each vaccine, according to the PRIME and UNIVAC models, are estimated as follows: USD 416,797 versus USD 416,801 for CECOLIN, USD 1,157,421 versus USD 1,154,245 for CERVARIX, USD 1,593,941 versus USD 1,588,890 for GARDASIL-4, and USD 2,812,605 versus USD 2,802,320 for GARDASIL-9 (see Figure 5 and Table 3).

**Figure 5:**
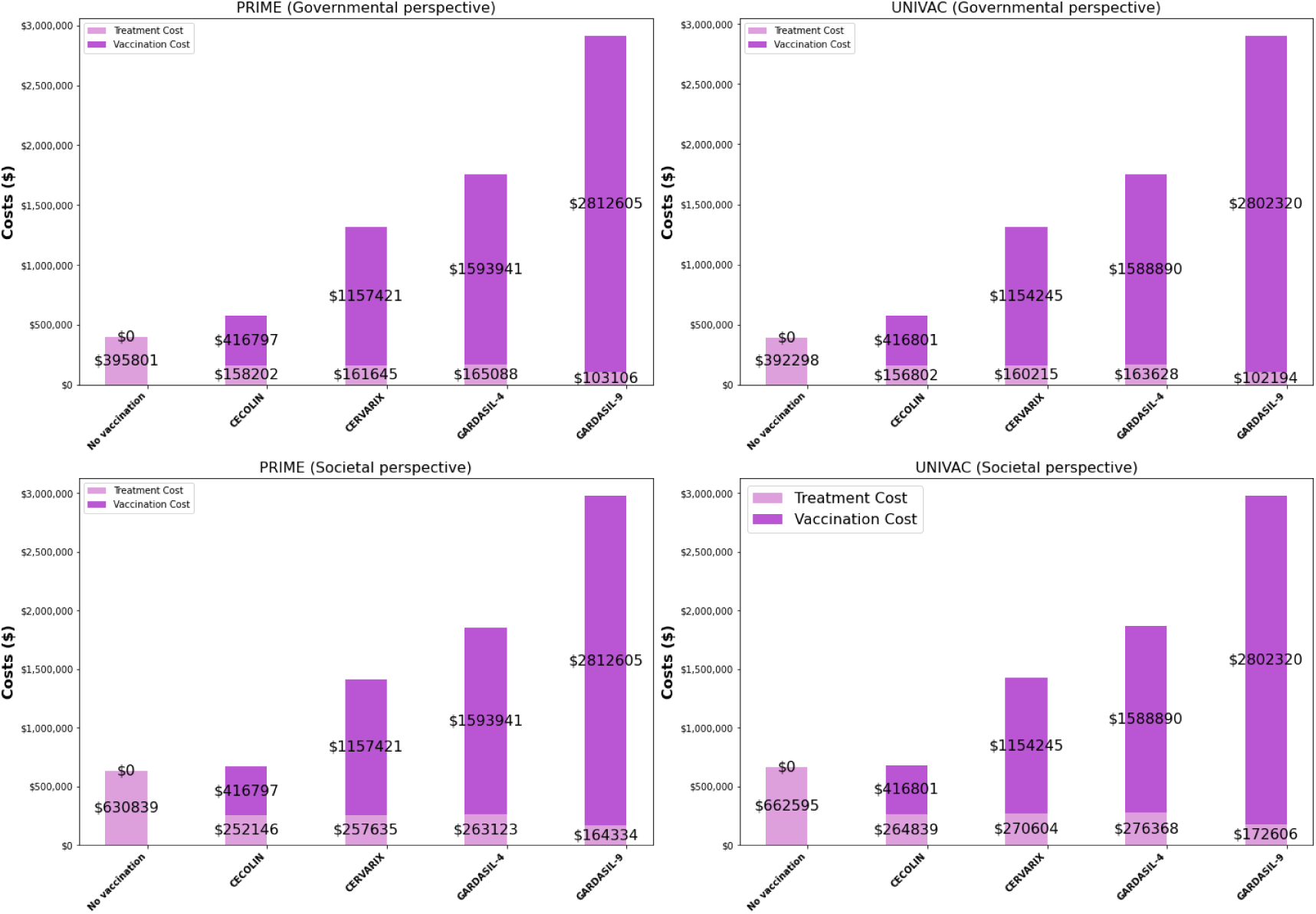
Total discounted economic costs associated with one-dose human papillomavirus (HPV) vaccination at 87 % coverage, over the lifetime of girls vaccinated at age 12 years alive in 2025. Vaccine program costs are estimated for each scenario of vaccination (CECOLIN, CERVARIX, GARDASIL-4 and GARDASIL-9). Healthcare treatment costs reflect disease costs associated with each strategy from a government and societal perspective (no cross protection).

**Table 3:** Discounted costs of HPV vaccination compared to no vaccination for CECOLIN and CERVARIX from government and societal perspectives. Lifetime economic outcomes using PRIME and UNIVAC models with cross protection.

| Economic outcomes | PRIME |  |  | UNIVAC |  |  |
| --- | --- | --- | --- | --- | --- | --- |
|  | No Vaccination | Vaccination | Difference | No Vaccination | Vaccination | Difference |
| **CECOLIN** |  |  |  |  |  |  |
| Vaccine program costs | 0 | 416797 | 416797 | 0 | 416801 | 416801 |
| Government healthcare costs | 395800 | 151315 | -244486 | 392298 | 149976 | -242323 |
| Societal healthcare costs | 630839 | 241170 | -389670 | 662595 | 253310 | -409285 |
| **CERVARIX** |  |  |  |  |  |  |
| Vaccine program costs | 0 | 1157421 | 1157421 | 0 | 993271 | 993271 |
| Government healthcare costs | 395800 | 120323 | -275477 | 392298 | 119259 | -273040 |
| Societal healthcare costs | 630839 | 191775 | -439064 | 662595 | 201429 | -461166 |

From a governmental perspective, and assuming no cross-protection, the three vaccines (CECOLIN, CERVARIX, and GARDASIL-4) are expected to avert approximately USD 235,000 in healthcare costs. In contrast, GARDASIL-9 is projected to avert USD 293,000 in healthcare costs in both the PRIME and UNIVAC models. Compared to no HPV vaccination, the averted healthcare costs for the three vaccines represent 60%, 59%, and 58% of the base case treatment costs, respectively. GARDASIL-9 vaccination program, due to its higher effectiveness in preventing cancer cases, results in even lower overall disease-specific costs, with averted healthcare costs representing 74%. From a societal perspective, the healthcare costs averted by CECOLIN, CERVARIX, and GARDASIL-4 are also around 60% of the base case costs, while for GARDASIL-9, this figure is 74% (see Figure 5). When considering cross-protection, CECOLIN is projected to avert 62% of healthcare costs compared to no HPV vaccination, both from governmental and societal perspectives. For CERVARIX, the estimated healthcare costs averted rise to 70% (see Table 3).

When comparing economic outcomes between the PRIME and UNIVAC models, the results are closely aligned. Both models estimate similar costs for each vaccine, with only marginal differences in projected outcomes. For instance, the costs associated with implementing CECOLIN, CERVARIX, GARDASIL-4, and GARDASIL-9 differ by less than 1% between the two models. Additionally, both models project similar levels of healthcare costs averted and cancer cases prevented across the different vaccines, reinforcing the robustness of the findings across varying modeling approaches.

### 3.3 Cost-effectiveness analysis

National pre-adolescent HPV vaccination in Tunisia was projected to be cost-effective compared to no vaccination in all scenarios evaluated as the cost-effectiveness ratios were less than GDP per capita.

In PRIME, without cross protection, one-dose bivalent vaccination (CECOLIN) has the lowest estimated net cost and most favourable cost-effectiveness ratio (0.04 (PRIME) times the national GDP per capita in Tunisia). CECOLIN vaccination was projected to avert 1109 DALYs compared to no vaccination over the birth cohort. Vaccination under this scenario was expected to incur $ 179198 more in discounted costs from the government perspective and $ 38104 more from the societal perspective compared to no vaccination. Further, the incremental cost per DALY averted was USD 845 (government perspective) and 718 (societal perspective) for CERVARIX vaccine. If cross-protection was not considered, CERVARIX would be dominated by CECOLIN because CERVARIX would generate less impact at a higher net cost. GARDASIL-4 is dominated by CECOLIN and CERVARIX because it averts fewer DALYs and costs more than both of these options. The cost per DALY averted was estimated to be $1266 from the government perspective and $1139 per DALY averted from the societal perspective for this scenario. GARDASIL-9 could achieve more benefit than CECOLIN but would be substantially more expensive with incremental cost-effectiveness of US$ 1845 from the government perspective (49% the national GDP per capita) and US$ 1718 from the societal perspective (0.46 times the national GDP per capita) (Table 4). With cross protection, CECOLIN had less favourable net cost than CERVARIX (US$172311 versus US$ 881944) but CERVARIX achieved substantially more health impact (70% versus 62%). In addition, CECOLIN would have favourable incremental cost-effectiveness (US$ 151 per DALY averted, or 0.04 times the national GDP per capita) when compared directly to CERVARIX.

**Table 4:** Cost per DALY averted for each scenario (CECOLIN, CERVARIX, GARDASIL-4 and GARDASIL-9) compared to no vaccination (with and without cross protection).

| Healtheconomic outcomes | PRIME |  | UNIVAC |  |
| --- | --- | --- | --- | --- |
|  | Government Perspective | Societal Perspective | Government Perspective | Societal Perspective |
| **CECOLIN** |  |  |  |  |
| Discounted net costs (USD) | 179198 | 38104 | 181304 | 19045 |
| Discounted DALYs averted | 1109 | 1109 | 1498 | 1498 |
| ICER (Cost per DALY averted) | 162 | 34 | 121 | 13 |
| **CERVARIX** |  |  |  |  |
| Discounted net costs (USD) | 923266 | 784217 | 922161 | 762254 |
| Discounted DALYs averted | 1092 | 1092 | 1476 | 1476 |
| ICER (Cost per DALY averted) | 845 | 718 | 625 | 516 |
| **GARDASIL-4** |  |  |  |  |
| Discounted net costs (USD) | 1363229 | 1226225 | 1360219 | 1202663 |
| Discounted DALYs averted | 1076 | 1076 | 1454 | 1454 |
| ICER (Cost per DALY averted) | 1266 | 1139 | 935 | 827 |
| **GARDASIL-9** |  |  |  |  |
| Discounted net costs (USD) | 2519911 | 2346100 | 2512216 | 2312331 |
| Discounted DALYs averted | 1366 | 1366 | 1845 | 1845 |
| ICER (Cost per DALY averted) | 1845 | 1718 | 1362 | 1253 |
| **CECOLIN (*cross protection*)** |  |  |  |  |
| Discounted net costs (USD) | 172311 | 27127 | 174478 | 7516 |
| Discounted DALYs averted | 1141 | 1141 | 1541 | 1541 |
| ICER (Cost per DALY averted) | 151 | 24 | 113 | 5 |
| **CERVARIX (*cross protection*)** |  |  |  |  |
| Discounted net costs (USD) | 881944 | 718357 | 881205 | 693079 |
| Discounted DALYs averted | 1285 | 1285 | 1737 | 1737 |
| ICER (Cost per DALY averted) | 686 | 559 | 507 | 399 |

In scenrios without cross protection, UNIVAC estimates that the incremental cost per DALY averted was USD 121 for CECOLIN, USD 625 for CERVARIX, USD 935 for GARDASIL-4 and USD 1362 for GARDASIL-9 from a government perspective. From a societal perspective, the incremental cost per DALY averted was USD 13 for CECOLIN, USD 516 for CERVARIX, USD 827 for GARDASIL-4 and USD 1253 for GARDASIL-9. Overall, when compared to no vaccination, all four vaccines were cost-effective strategies for the prevention of cervical cancer. The incremental cost per DALY averted for all four vaccines was below the cost-effective threshold of USD 3747 (Tunisia’s GDP per capita). CECOLIN has the lowest net cost and most attractive cost-effectiveness ($121 per DALY averted from a government perspective and cost-saving from a societal perspective). CERVARIX and GARDASIL-4 would be dominated by CECOLIN because these options would generate less impact at a higher net cost. The incremental cost-effectiveness of the remaining alternative (GARDASIL-9) would exceed 0.3 times the national GDP per capita from either a government or societal perspective (Table 4). If cross protection was assumed, the estimated incremental cost per DALY averted by CECOLIN was lower than CERVARIX.

We estimate that all HPV vaccination in Tunisia will prevent a substantial number of cervical cancer cases and deaths. Dominated options are more expensive and generate fewer benefits than at least one alternative option. The efficiency frontier links the interventions that are not dominated and provides guidance. Any strategy that is placed on the frontier is reasonably efficient. Without cross-protection, CECOLIN is likely to be the preferred product, generating lower net costs and similar benefits to both GARDASIL-4 and CERVARIX. With cross-protection, CECOLIN also had the most favourable cost-effectiveness, even if CERVARIX generated substantially more health benefits than CECOLIN (70% versus 62% vaccine impact). As CERVARIX had less favourable incremental cost-effectiveness compared to CECOLIN (507/DALY versus 113/DALY), this option should be given serious consideration if affordable. Our findings also suggest that GARDASIL-9 is unlikely to be a viable option unless the assumed price per dose is substantially reduced (Table 4 and Fig. 6)

**Figure 6:**
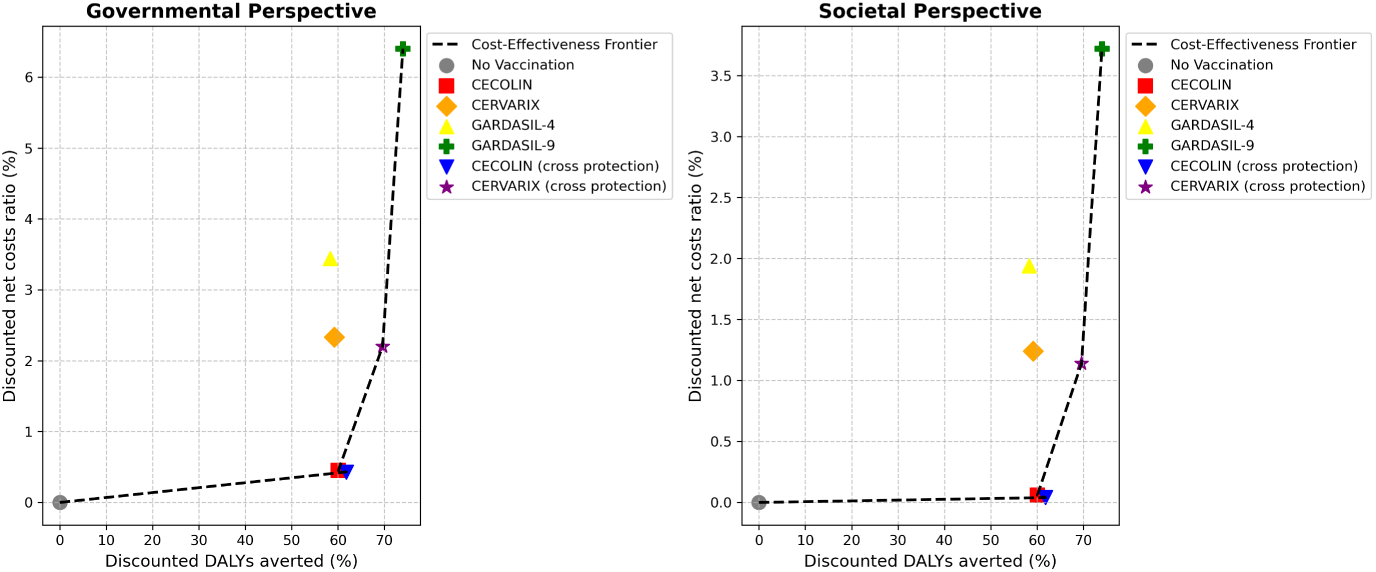
Cost-effectiveness frontier between total discounted costs per scenario and Ddiscounted DALYs. Discounted costs (vaccination and treatment) and DALYs are drawn for all the strategies assessed in the base case scenario. Strategies include vaccination with CECOLIN, CERVARIX, GARDASIL-4 and GARDASIL-9. Bivalent vaccines differ by assuming cross protection or not. Names of the strategies located on the cost-effectiveness frontier compared with the lower-cost non-dominated strategy are shown.

### 3.4 Sensitivity analysis

One-way deterministic sensitivity analysis was performed to evaluate the robustness of the results for the most cost-effective options (CECOLIN and CERVARIX with cross-protection) compared to no vaccination. Age of vaccination, cancer incidence rate, the proportion of cases that end in cervical cancer death, treatment cost from government perspective, and discount rate were varied to determine the effect of uncertainty on the results of incremental cost per DALY averted (Table 5) from the government perspective. The sensitivity results show that the annual discount rate for future benefits and costs, and disease burden rates tended to have the most influence on model outcomes. Other variables, such as the target age group and healthcare treatment costs, were less influential on cost-effectiveness results; across reasonable ranges of values for these parameters, the qualitative results generally remained consistent (results are provided in figure 9 in the Appendix 5). Across scenarios and perspectives, when varying individual parameters for uncertainty impacts, the cost per DALY averted for vaccination among girls ranged from cost-saving to $13114. This represents up to three times the Tunisia’s GDP per capita of $3895.4 (USD$ 2023). Adjusting the discount rate by +7% caused the biggest change in the ICER value and may change the conclusions. Under this scenario, the cost per DALY averted was equivalent to 1.32 times (USD 5154) and 3.37 times (USD 13114) of the GDP per capita for CECOLIN and CERVARIX with cross protection. The model results were robust, and the discount rate was the main factor affecting the baseline analysis.

**Table 5:**
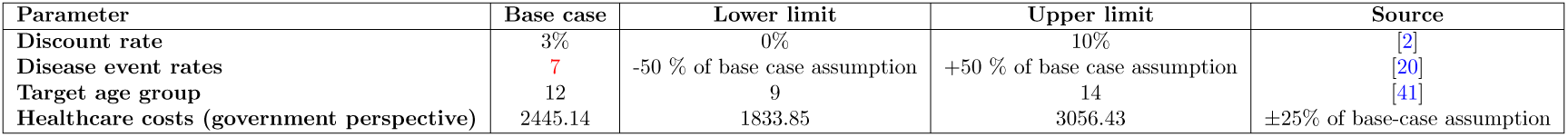
Input parameters for univariate sensitivity analysis.

| Parameter | Base case | Lower limit | Upper limit | Source |
| --- | --- | --- | --- | --- |
| Discount rate | 3% | 0% | 10% | [2] |
| Disease event rates | 7 | -50 % of base case assumption | +50 % of base case assumption | [20] |
| Target age group | 12 | 9 | 14 | [41] |
| Healthcare costs (government perspective) | 2445.14 | 1833.85 | 3056.43 | $\pm 25\%$ of base-case assumption |

Tunisia, like many countries, does not have established cost-effectiveness thresholds for health interventions, including vaccination. However, results from the probabilistic sensitivity analyses determined 100% credible ranges around the ratios for each of the five cost-effective vaccination scenarios (CECOLIN, CERVARIX, with and without cross protection and GARDASIL4) at a willingness-to-pay threshold (WTP) of $1169 per DALY averted (which corresponds to 30% GDP per capita) from the government and the societal perspectives (fig. 7b and 8b).

**Figure 7:**
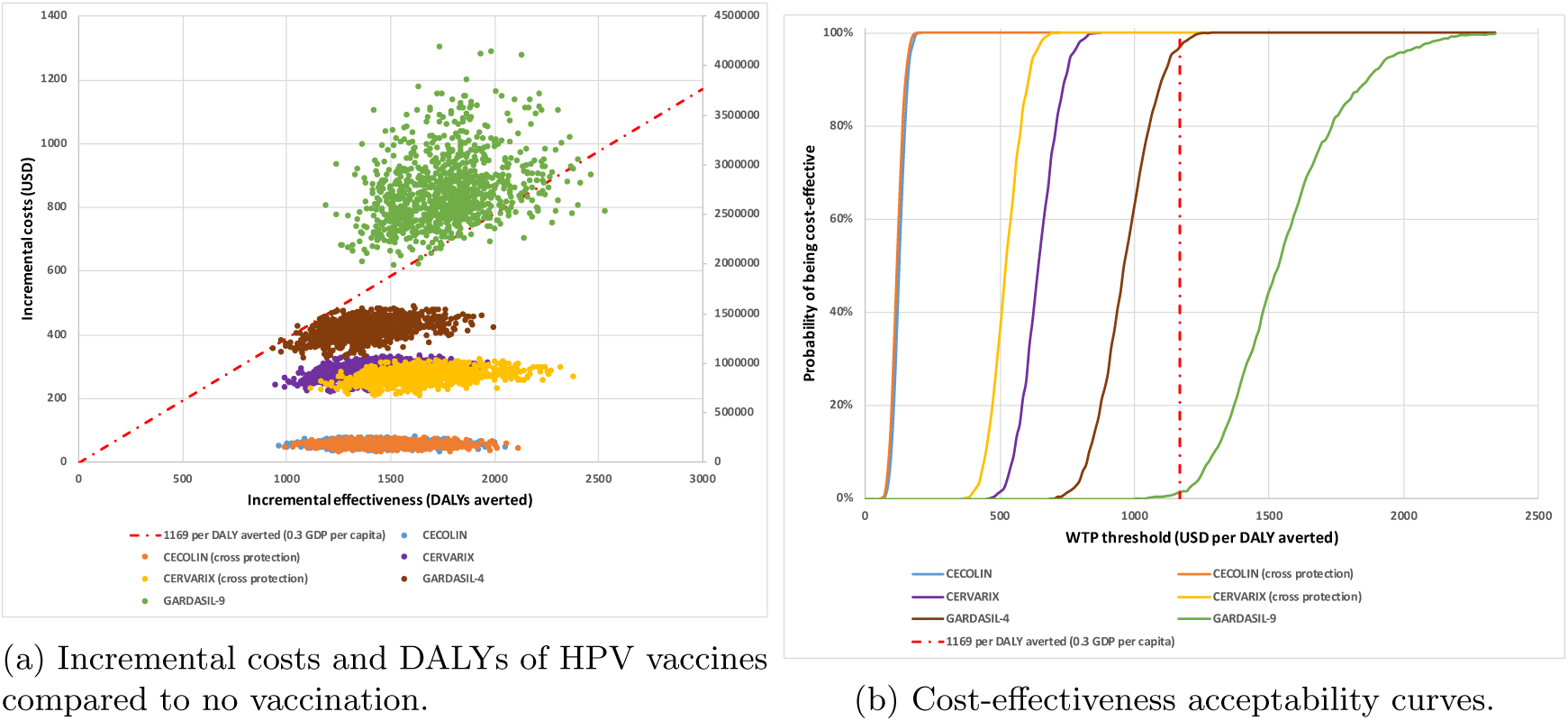
Probabilistic clouds showing the incremental costs (USD) and effectiveness (DALYs averted) of each HPV vaccine product (CECOLIN, CERVARIX, GARDASIL-4 and GARDASIL-9) without cross protection and with cross protection for the favourable cost-effective vaccines (CECOLIN and CERVARIX), compared to no vaccine. The cost-effectiveness acceptability curves (right) demonstrate likelihood of vaccines cost-effectiveness across varying willingness to pay thresholds (government perspective).

**Figure 8:**
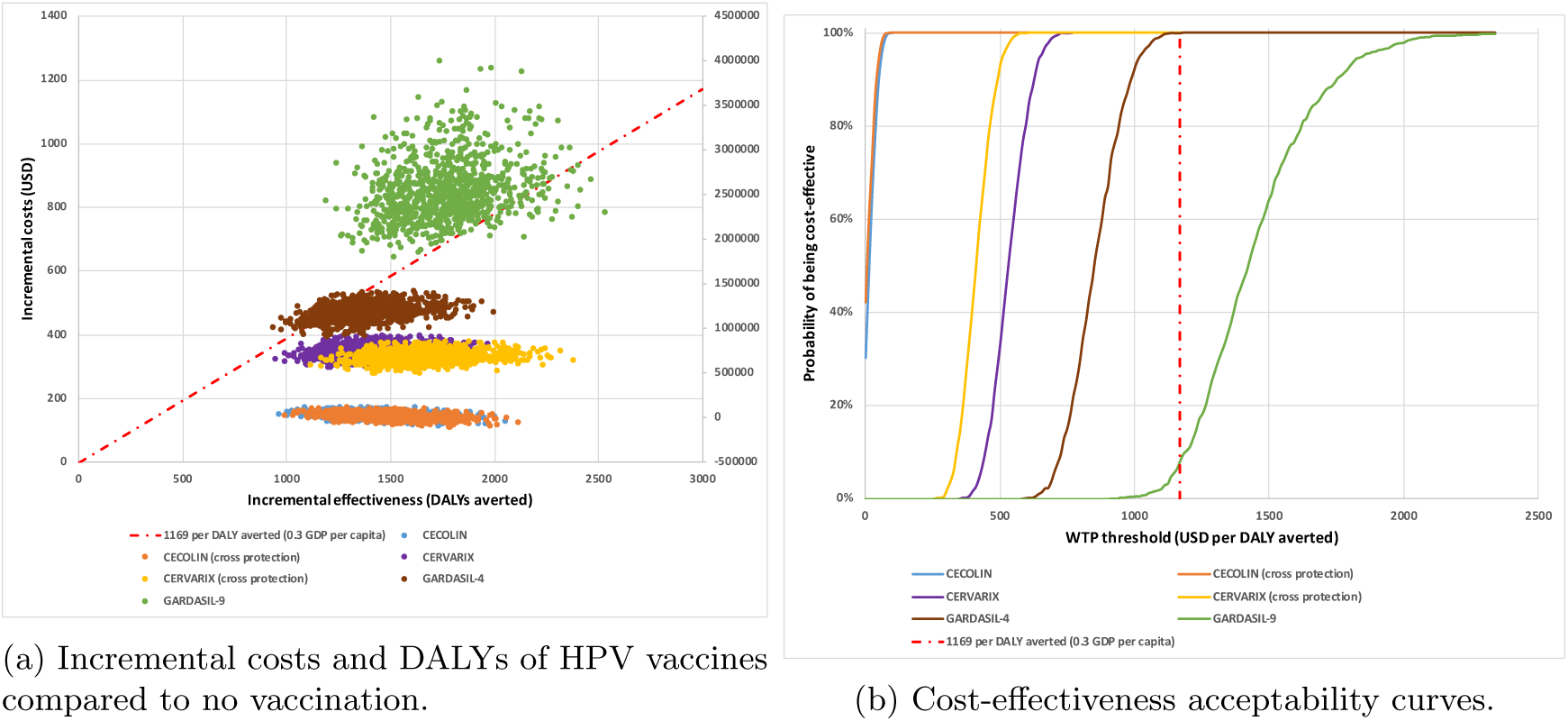
Probabilistic clouds showing the incremental costs (USD) and effectiveness (DALYs averted) of each HPV vaccine product (CECOLIN, CERVARIX, GARDASIL-4 and GARDASIL-9) without cross protection and with cross protection for the favourable cost-effective vaccines (CECOLIN and CERVARIX), compared to no vaccine. The cost-effectiveness acceptability curves (right) demonstrate likelihood of vaccines cost-effectiveness across varying willingness to pay thresholds (societal perspective).

Without cross protection, CECOLIN and CERVARIX had a similar probability of being cost-effective compared to no vaccination and probabilistic uncertainty clouds associated overlap from both a government and societal perspective (fig. 7 and 8). CECOLIN had the most favorable cost-effectiveness, but GARDASIL-9 provided greater health benefits given the available alternatives and could also be considered if affordable (less than 5% probability that would be cost-effective at a threshold set at around US$ 1169 the national GDP per capita). It should be noted that vaccine prices were fixed for the probabilistic sensitivity analysis and varied for GARDASIL-9 only, therefore, the relative position of the probabilistic clouds will be very sensitive to changes in other vaccine prices.

However, with cross-protection, comparing the products with the most favorable cost-effectiveness (CECOLIN and CERVARIX with cross-protection) had a similar 100 % probability of being cost-effective at a WTP threshold set at $600 (15 % of Tunisia’s national GDP per capita) and $ 700 (18 % of GDP) when compared to no vaccination from a government and societal perspective, respectively (figure 7b and 8b). However, CECOLIN is a far more attractive option with the most favorable cost-effectiveness as it would have a 100 % probability of being cost-effective at a WTP threshold set at $ 200 (5 % of Tunisia’s national GDP per capita) when compared to no vaccination.

## 4 Discussion

We assessed the long-term cost-effectiveness of implementing HPV vaccination for 12-year-old girls in Tunisia by 2025. Our results indicate that the vaccination could decrease the incidence of cervical cancer cases and related deaths by 58–74%, depending on assumptions regarding cross-protection. Moreover, the introduction of HPV vaccination is projected to be cost-effective in all scenarios considered. We find that the most cost-effective vaccine would be either cost-saving or cost-effective at a WTP threshold set at 30% GDP per capita. Our results were particularly sensitive to the choice of vaccine product, cross-protection assumptions, vaccine price, and discount rate because the benefits of HPV vaccination occur many years in the future. For instance, assigning a higher discount rate is, therefore, unfavourable to HPV vaccination.

Our estimates of the cost-effectiveness of HPV vaccination are similar to other estimates for Tunisia presented as part of health and economic evaluation analyses in the literature. A study by [20] on the model-based impact and cost-effectiveness of cervical cancer prevention in the extended middle east and north africa (EMENA) estimated a cost per DALY averted of 100-$1400 in Tunisia (USD 2012). A second study by Jit et al evaluating vaccine cost-effectiveness in 179 countries projected a cost per DALY averted with HPV vaccination of 597 for vaccinating 12 year old girls (USD 2014) [3]. A third study by Messoudi et al. (2018) [23] on cost-effectiveness of HPV vaccine introduction in Morocco for girls aged 14 years old concluded that vaccination alone was the most cost-effective strategy with an ICER of USD 207 per years of life saved. Furthermore, other studies have demonstrated the substantial public health benefits of vaccinating young girls, showing that such programs can prevent numerous cases of cervical cancer and save lives [4, 21, 26, 33, 1] (Kenya, Ghana, Mozambique, Burkina Fao and Ethiopia). These findings are in line with the cost-effectiveness ratios estimated in our study for each scenario: 151 $ (PRIME) and 24 $ (UNIVAC) per DALY averted from the government perspective and 113 $ (PRIME) and 5 $ (UNIVAC) per DALY averted from the societal perspective for CECOLIN assuming cross protection.

## 5 Conclusion

HPV vaccination is a cost-effective intervention in Tunisia. The optimal choice of vaccine depends on influential assumptions about cross-protection. The cost-effectiveness of the vaccines should be continually re-evaluated as more information emerges about their efficacy and costs.

Our study had some limitations. First, UNIVAC and PRIME are static cohort models and therefore, not captures any additional indirect (herd immunity) benefits associated with vaccination. However, these effects would only have made our results more favourable to vaccination. Second, we excluded costs borne by households, such as out-of-pocket medical expenses and lost earnings. However, these costs are likely to be relatively small, and a preliminary analysis with these costs included did not alter the cost-effectiveness results. Additionally, the model does not account for the costs or disease burden associated with the prevention, detection, or treatment of pre-cancerous lesions, which are significant contributors to the overall burden of cervical cancer. However, Tunisia lacks a national cervical cancer screening program, and current screening rates are low.

Evidence indicates that without a substantial and immediate expansion of vaccination, screening, and treatment efforts, cervical cancer-related deaths in low- and middle-income countries (LMICs) could increase by up to 50% by 2040 [24]. In Tunisia, introducing a national HPV vaccination program for girls would be a highly cost-effective measure to significantly reduce the burden of cervical cancer. However, the economic benefits of vaccination must be carefully weighed alongside considerations of budget impact, affordability, feasibility, equity, and other local factors to ensure the successful and sustainable integration of the vaccine into the national health strategy.

## Data Availability

All data produced in the present work are contained in the manuscript

## Acknowledgements

This work was supported, in part, by Vaccine Impact Modelling Consortium (VIMC) and by the Bill & Melinda Gates Foundation [INV-059607]. Under the grant conditions of the Foundation, a Creative Commons Attribution 4.0 Generic License has already been assigned to the Author Accepted Manuscript version that might arise from this submission. At the time of analysis, the VIMC was jointly funded by Gavi, the Vaccine Alliance and the Bill & Melinda Gates Foundation (grant numbers INV-034281 and INV-009125/OPP1157270).

## A Appendix 1: Model comparaison

Both the PRIME and UNIVAC models are static multi-cohort, proportional impact models used to estimate the impact of HPV vaccination on cervical cancer cases and deaths. They maintain constant age-specific cervical cancer incidence among unvaccinated women and assume conservative estimates without cross-protection or indirect effects. The UNIVAC model evaluates catch-up campaigns, stratified cervical cancer cases by stage, and hospitalizations. In contrast, the PRIME model, supported by WHO, focuses on the cost-effectiveness of vaccinating females before sexual debut, utilizing country-specific data and customizable inputs for vaccine efficacy, price, delivery, and cancer treatment costs.

## B Appendix 2: Inputs for disease burden

## C Appendix 3: Input parameters for health service costs from the government and societal perspective

## D Appendix 4: Vaccine program costs

## E Appendix 5: Deterministic sensitivity analysis

**Table 6:**
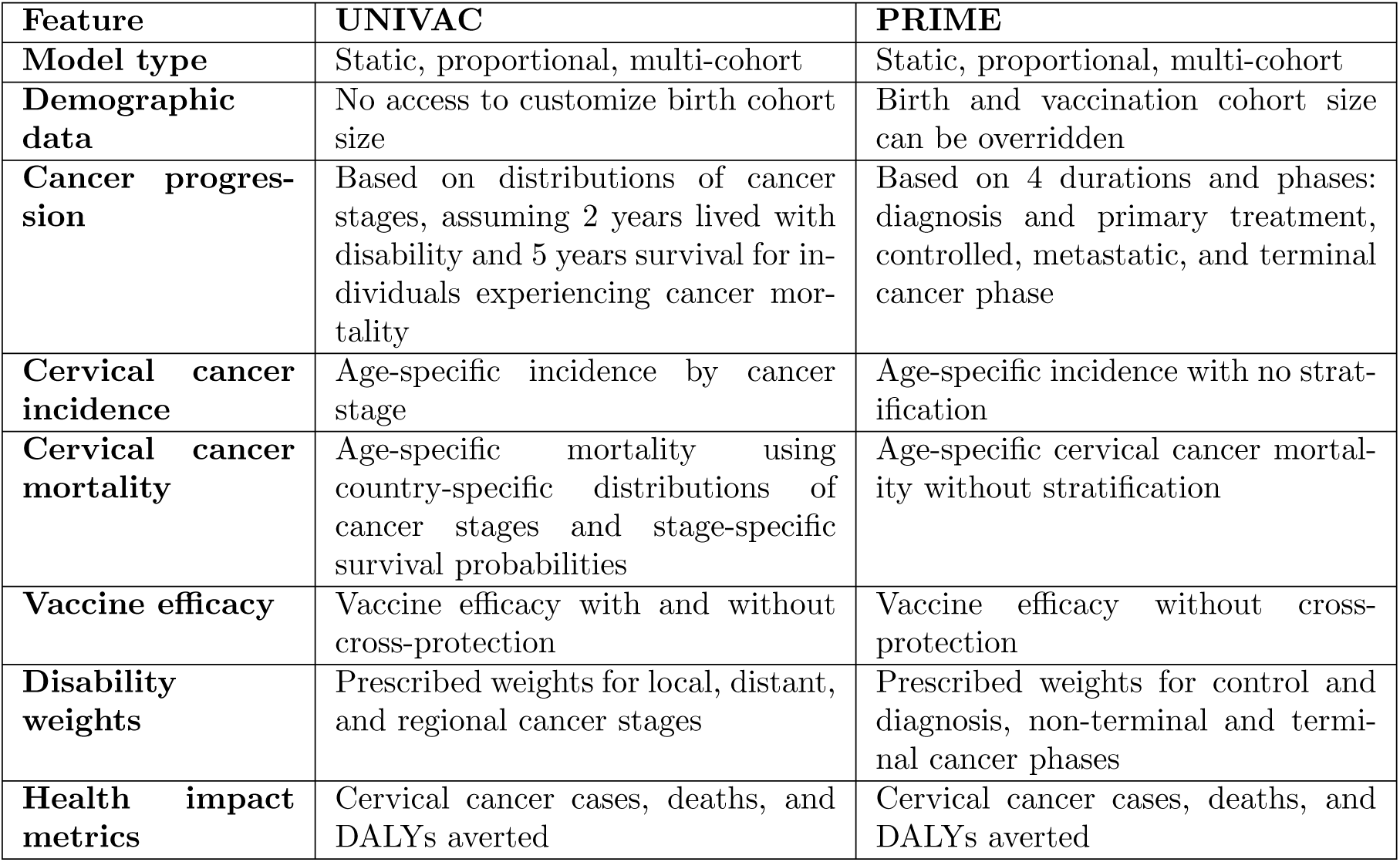
Overview of PRIME and UNIVAC models for cost-effectiveness of HPV vaccination.

**Table 7:**
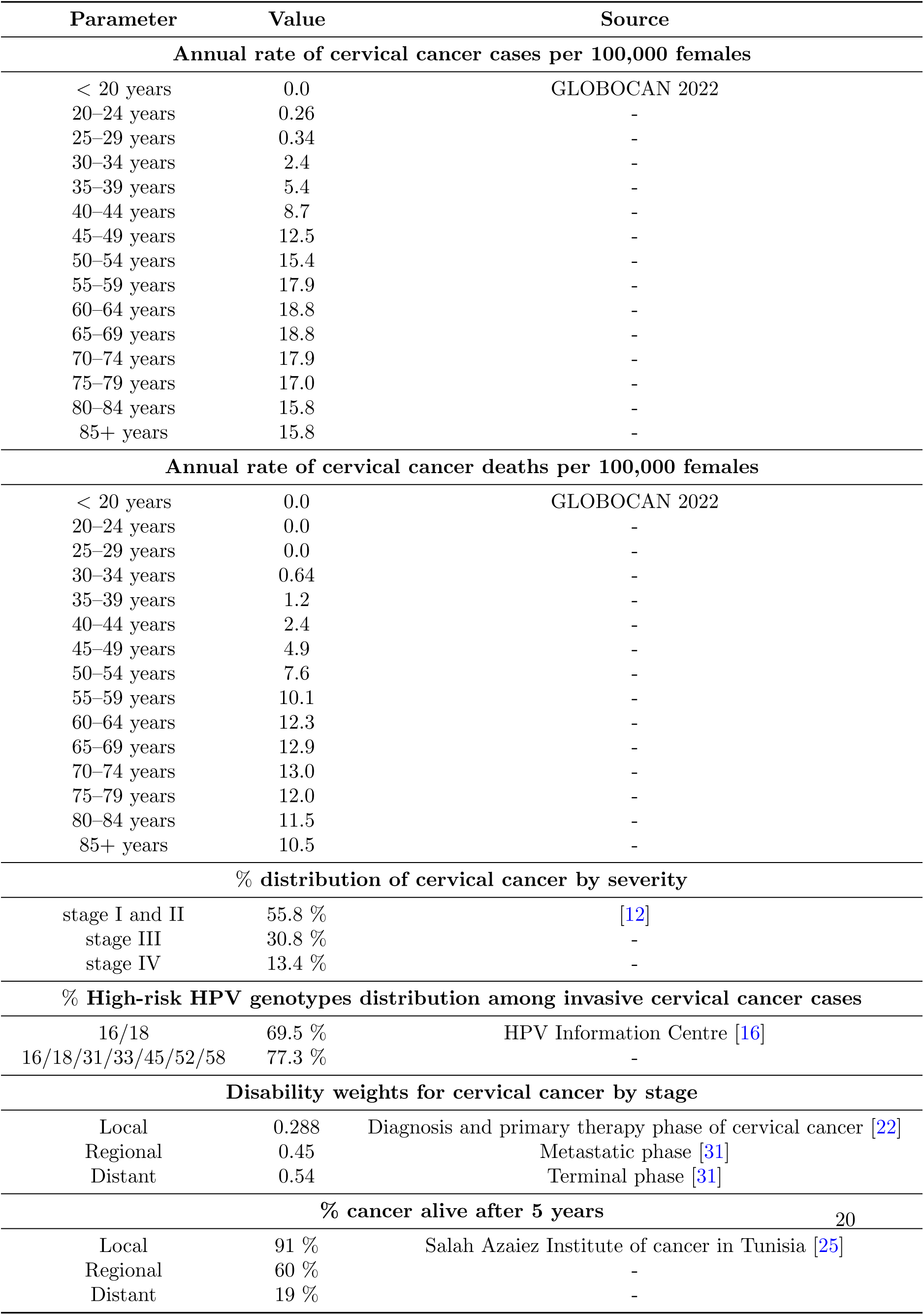
Input parameters for estimating cervical cancer disease burden.

**Table 8:** Average cost per treated case of cervical cancer (USD).

| Parameter | Value | Source |
| --- | --- | --- |
| **CC treatment cost per case (government perspective)** |  |  |
| Local | 2603 | Salah Azaiez Institute of cancer in Tunisia [25] |
| Distant | 1800 | - |
| Regional | 750 | - |
| Average cancer treatment cost per case | 2445.14 | - |
| **CC treatment cost per case (societal perspective)** |  |  |
| Local | 4055 | [35, 25] |
| Distant | 3252 | - |
| Regional | 2202 | - |
| Average cancer treatment cost per case | 3897,14 | - |

**Table 9:**
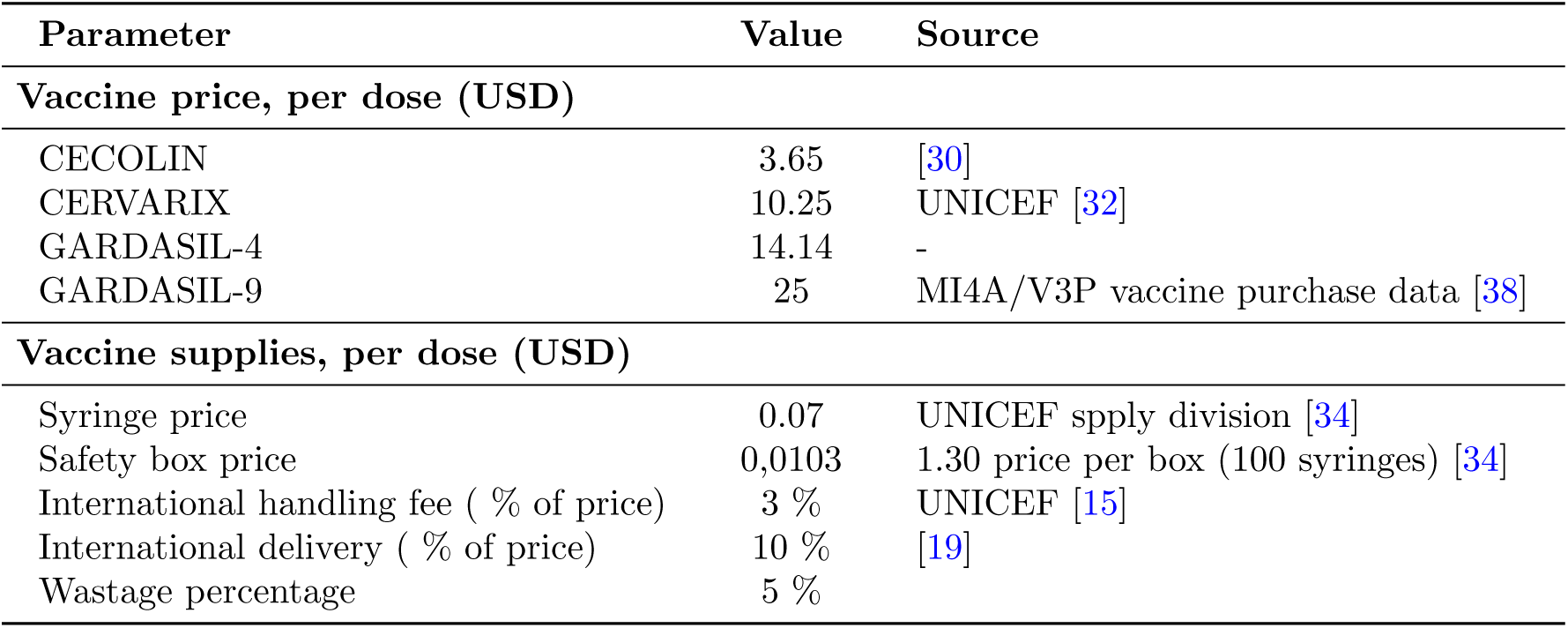
Input parameters for estimating HPV vaccine program costs.

**Figure 9:**
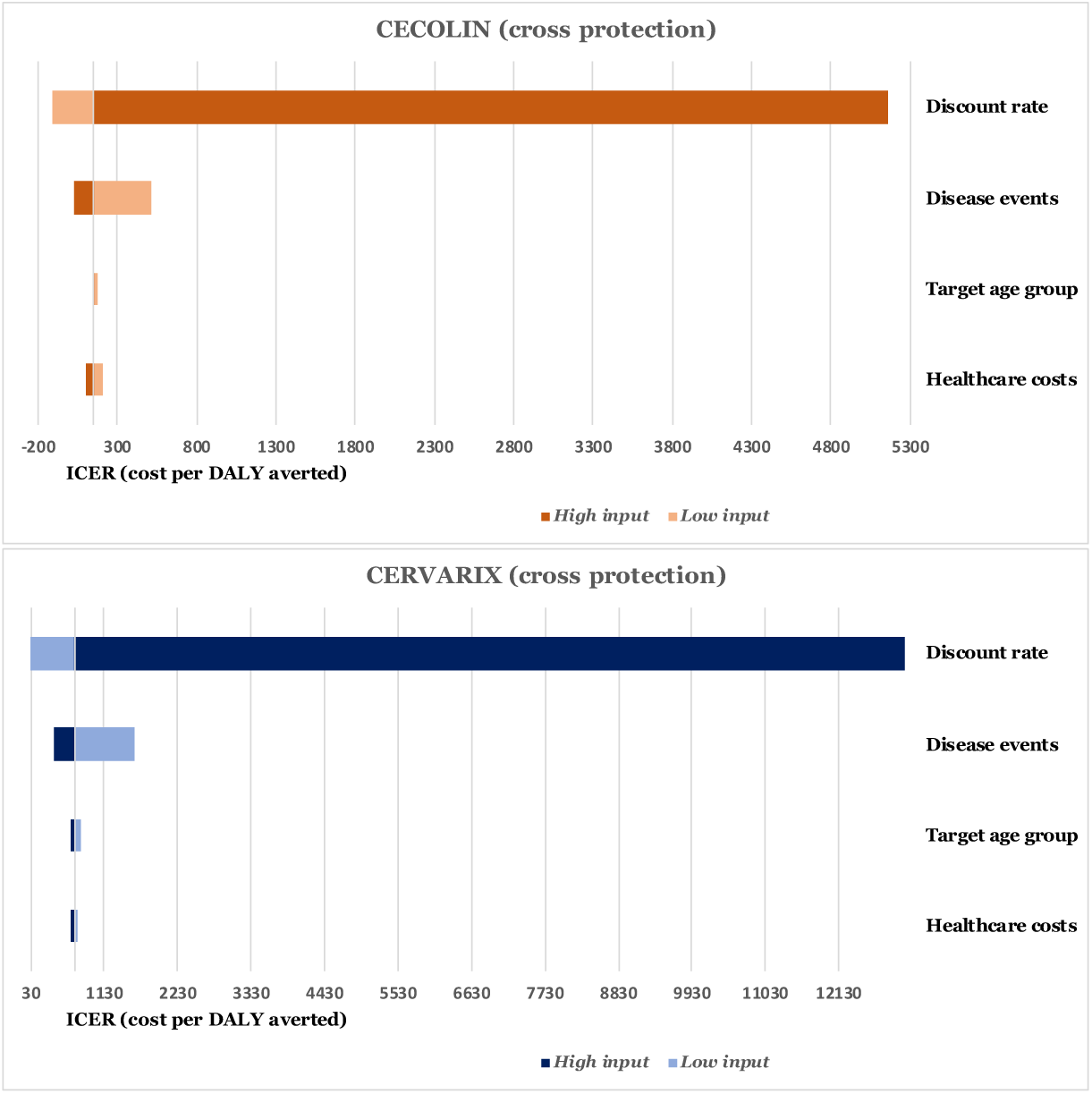
Univariate sensitivity analysis for CECOLIN and CERVARIX assuming cross protection.

## References

[1] Wondimu A, Postma MJ, van Hulst M. Cost-effectiveness analysis of quadrivalent and nonavalent human papillomavirus vaccines in Ethiopia. Vaccine. 2022 Mar 25;40(14):2161–2167. doi: 10.1016/j.vaccine.2022.02.080. Epub 2022 Mar 2. PMID: 35248423.

[2] WHO. WHO guide for standardization of economic evaluations of immunization programmes;2019. https://iris.who.int/bitstream/handle/10665/329389/WHO-IVB-19.10-eng.pdf?sequence=1. Accessed 24 July 2024.

[3] Jit M, Brisson M, Portnoy A, Hutubessy R. Cost-effectiveness of female human papillomavirus vaccination in 179 countries: a PRIME modelling study. Lancet Glob Health. 2014;2(7):e406–14.

[4] Mwenda V, Jalang’o R, Miano C, Bor JP, Nyangasi M, Mecca L, Were V, Kariithi E, Pecenka C, Schuind A, Abbas K, Clark A. Impact, cost-effectiveness, and budget implications of HPV vaccination in Kenya: A modelling study. Vaccine. 2023 Jun 29;41(29):4228–4238. doi: 10.1016/j.vaccine.2023.05.019. Epub 2023 Jun 8. PMID: 37296015.

[5] Brown, D. R., Joura, E. A., Yen, G. P., Kothari, S., Luxembourg, A., Saah, A., Walia, A., Perez, G., Khoury, H., Badgley, D., & Stanley, M. (2021). Systematic literature review of cross-protective effect of HPV vaccines based on data from randomized clinical trials and real-world evidence. *Vaccine*, 39(7), A1–A12. 10.1016/j.vaccine.2021.03.024

[6] FUTURE II Study Group. Quadrivalent vaccine against human papillomavirus to prevent high-grade cervical lesions. N Engl J Med. 2007;356(19):1915–1927.

[7] Joura EA, Giuliano AR, Iversen OE, et al. A 9-valent HPV vaccine against infection and intraepithelial neoplasia in women. N Engl J Med. 2015;372(8):711–723.

[8] Chan, C.K.; Aimagambetova, G.; Ukybassova, T.; Kongrtay, K.; Azizan, A. Human Papillomavirus Infection and Cervical Cancer: Epidemiology, Screening, and Vaccination-Review of Current Perspectives. J. Oncol. 2019, 2019, 3257939.

[9] de Sanjose S, Quint WG, Alemany L, et al. Human papillomavirus genotype attribution in invasive cervical cancer: a retrospective cross-sectional worldwide study. Lancet Oncol 2010;11:1048–56.

[10] WHO. One-Dose Human Papillomavirus (HPV) Vaccine Offers Solid Protection against Cervical Cancer. 2022. Available online: https://www.who.int/news/item/11-04-2022-one-dose-human-papillomavirus-(hpv)-vaccine-offers-solid-protection-against-cervical-cancer.

[11] PAHO. PROVAC Toolkit. Available from: https://www.paho.org/provac-toolkit/tools/about-univac/; n.d.

[12] M. Garci, Cancer du col de l’utérus : étude épidémiologique multicentrique, Thèse de doctorat en Médecine, Université de Tunis El Manar, Faculté de Médecine de Tunis, 2020.

[13] International Agency for Research on Cancer (IARC). Cancer Today [Internet] [accessed 2024 July 29]. Available from: https://gco.iarc.fr/today/home.

[14] Barnabas RV, Brown ER, Onono MA, Bukusi EA, Njoroge B, Winer RL, et al. Efficacy of Single-Dose Human Papillomavirus Vaccination among Young African Women. NEJM Evid 2022 Apr 11;1(5).

[15] UNICEF Supply Division. Handling fees [Internet]. [accessed 2024 July 31]. Available from: https://www.unicef.org/supply/handling-fees.

[16] HPV Information Centre [Internet]. [accessed 2024 July 29]. Available from: https://hpvcentre.net/index.php.

[17] Vodicka E, Nonvignon J, Antwi-Agyei KO, Bawa J, Clark A, Pecenka C, et al. The projected cost-effectiveness and budget impact of HPV vaccine introduction in Ghana. Vaccine. 2022;40:A85–93.

[18] PaavonenJ,nJenkinsD,nBoschFX, et al. Efficacy of aprophylactic adjuvanted bivalent L1 virus-like-particle vaccine against infection with human papillomavirus types 16 and 18 in young women: an interim analysis of a phase III double-blind, randomised controlled trial. Lancet. 2007;369(9580):2161-2170.

[19] Vodicka E, Nonvignon J, Antwi-Agyei KO, Bawa J, Clark A, Pecenka C, et al. The projected cost-effectiveness and budget impact of HPV vaccine introduction in Ghana. Vaccine 2022 Mar;31(40):A85–93.

[20] Kim JJ, Sharma M, O’Shea M, Sweet S, Diaz M, Sancho-Garnier H, Seoud M. Model-based impact and cost-effectiveness of cervical cancer prevention in the Extended Middle East and North Africa (EMENA). Vaccine. 2013 Dec 30;31 Suppl 6:G65–77. doi: 10.1016/j.vaccine.2012.06.096. PMID: 24331822.

[21] Vodicka E, Nonvignon J, Antwi-Agyei KO, Bawa J, Clark A, Pecenka C, LaMontagne DS. The projected cost-effectiveness and budget impact of HPV vaccine introduction in Ghana. Vaccine. 2022 Mar 31;40 Suppl 1:A85–A93. doi: 10.1016/j.vaccine.2021.07.027. Epub 2021 Jul 21. PMID: 34303563.

[22] Abbas KM, van Zandvoort K, Brisson M, Jit M. Effects of updated demography, disability weights, and cervical cancer burden on estimates of human papillomavirus vaccination impact at the global, regional, and national levels: a PRIME modelling study. The Lancet Global Health. 2020;8(4):e536–e544.

[23] Messoudi, W., El Mahi, T., Diaz Sanchiz, M., Saadani, G., Zidouh, A., Nejjari, C., & Tachfouti, N. (2018). Cost-effectiveness of HPV vaccine introduction in Morocco. Revue d’Épidémiologie et de Santé Publique, 66(S1), S101. 10.1016/j.respe.2018.03.109.

[24] Qiao YL, Wu T, Li RC, Hu YM, Wei LH, Li CG, et al. Efficacy, safety, and immunogenicity of an escherichia coli-produced bivalent human papillomavirus vaccine: An interim analysis of a randomized clinical trial. J Natl Cancer Inst 2020;112(2):145–53.

[25] E. Mziou, H. Khiari, M. Gargouri, M. Hsairi, Stage-specific treatment costs for cervical cancer in the Salah Azaiez Institute of cancer in Tunisia, Biomedicine & Healthcare Research, Vol. 2 No. Issue 2, 2024. 10.1016/j.jcpo.2020.100237.

[26] Kiendrébéogo JA, Sidibe ARO, Compaoré GB, Nacanabo R, Sory O, Ouédraogo I, Nawaz S, Schuind AE, Clark A. Cost-effectiveness of human papillomavirus (HPV) vaccination in Burkina Faso: a modelling study. BMC Health Serv Res. 2023 Dec 1;23(1):1338. doi: 10.1186/s12913-023-10283-3. PMID: 38041075; PMCID: PMC10693094.

[27] National Institute of Statistics Tunisia. [Internet]. [cited 2024 Aug 13]. Available from: http://www.ins.tn/en.

[28] Dasbach EJ, Elbasha EH, Insinga RP. Mathematical models for predicting the epidemiologic and economic impact of vaccination against human papillomavirus infection and disease. Epidemiol Rev. 2006;28:88–100. doi: 10.1093/epirev/mxj006. Epub 2006 Jun 1. PMID: 16740585.

[29] Ryser MD, Gravitt PE, Myers ER. Mechanistic mathematical models: An underused platform for HPV research. Papillomavirus Res. 2017 Jun;3:46–49. doi: 10.1016/j.pvr.2017.01.004. Epub 2017 Feb 4. PMID: 28720456; PMCID: PMC5518640.

[30] Ministry of Health. Tunisia Health Information System, unpublished data; 2024.

[31] Salomon JA, Haagsma JA, Davis A, de Noordhout CM, Polinder S, Havelaar AH. Disability weights for the Global Burden of Disease 2013 study. Lancet Global Health 2015:e712–23.

[32] UNICEF. Human Papilloma Virus (HPV) vaccine price data [Internet]. [accessed 2024 July 31]. Available from: https://www.unicef.org/supply/documents/human-papilloma-virus-hpv-vaccine-price-data.

[33] Guimaräes EL, Chissaque A, Pecenka C, Debellut F, Schuind A, Vaz B, Banze A, Rangeiro R, Mariano A, Lorenzoni C, Carrilho C, Martins MDRO, de Deus N, Clark A. Impact and Cost-Effectiveness of Alternative Human Papillomavirus Vaccines for Preadolescent Girls in Mozambique: A Modelling Study. Vaccines (Basel). 2023 Jun 2;11(6):1058. doi: 10.3390/vaccines11061058. PMID: 37376447; PMCID: PMC10304296.

[34] UNICEF. Syringe and safety box bundles price data [Internet]. [accessed 2024 July 31]. https://www.unicef.org/supply/documents/syringe-and-safety-box-bundles-price-data.

[35] Van Minh H, My NTT, Jit M. Cervical cancer treatment costs and cost-effectiveness analysis of human papillomavirus vaccination in Vietnam: a PRIME modeling study. BMC Health Serv Res. 2017;17(1):353.

[36] UNICEF Supply Division. EPI Logistics Forecasting Tool 2022 [spreadsheet].

[37] World Health Organization. Global strategy to accelerate the elimination of cervical cancer as a public health problem. Geneva: World Health Organization, 2020.

[38] World Health Organization. Immunization, Vaccines and Biologicals. MI4A Vaccine purchase data [Internet]. [accessed 2024 July 31]. Available from: https://www.who.int/teams/immunization-vaccines-and-biologicals/vaccine-access/mi4a/mi4a-vaccine-purchase-data.

[39] Wheeler CM, Castellsagué X, Garland SM, Szarewski A, Paavonen J, Naud P, et al. Cross-protective efficacy of HPV-16/18 AS04-adjuvanted vaccine against cervical infection and precancer caused by non-vaccine oncogenic HPV types: 4-year end-of-study analysis of the randomised, double-blind PATRICIA trial. Lancet Oncol.

[40] Wahl B, Gupta M, Erchick DJ, et al. Change in full immunization inequalities in Indian children 12–23 months: an analysis of household survey data. BMC Public Health. 2021;21(1):841.

[41] World Health Organization. Global strategy to accelerate the elimination of cervical cancer as a public health problem. Geneva: World Health Organization; 2020.

